# Mechanisms of increased Alzheimer’s Disease pathology with *R47H* and *R62H TREM2* variants

**DOI:** 10.1101/2022.07.12.22277509

**Authors:** Nurun Fancy, Nanet Willumsen, Vicky MN Chau, Samuel L Boulger, Harry J. Whitwell, Wenhao Wang, Baptiste Avot, Michael Thomas, Jonathan Talbot-Martin, Stergios Tsartsalis, Combiz Khozoie, Aisling McGarry, Eleonore Schneegans, Riad Yagoubi, Dorcas Cheung, Marianna Papageorgopoulou, Emily Adair, Benjamin Cooper, Karen Davey, Amy M Smith, William Scotton, John Hardy, Paul M Matthews, Johanna S Jackson

**Author notes:** joint senior authors (Paul M Matthews, Johanna S Jackson). Corresponding authors, 020 7594 2241, 020 7594 2855.

## Abstract

*TREM2* plays multiple functional roles in microglia and variants are associated with increased risks of Alzheimer’s disease (AD). Genetic polymorphisms reducing expression of the functionally related protein CD33 are protective. Here we have contrasted cellular pathology in human *post-mortem* brain with and without AD to test mechanisms associated with the differential genetic risks conferred by *R47H* and *R62H TREM2* variants (*TREM2var*) with and without heterozygosity for the protective rs3865444 *CD33* polymorphism. Epistasis between *CD33* and *TREM2* was demonstrated by both relative normalisation of differences in β-amyloid load in *TREM2var* carriers of the protective *CD33* allele. These functional differences were mirrored by differential microglial transcriptomic responses to β-amyloid. Controlling for *CD33* genotype, microglial transcriptional responses to increasing β-amyloid were lower for *TREM2var*, particularly for *R47H* compared to *CV* and there was a reduction in expression of neuroplasticity pathways in *TREM2var. R62H* microglial signatures were distinguished from those of *R47H* by upregulation of genes associated with phagocytosis and from *CV* by differences in inflammatory gene expression including those involved in NK-kappaB signalling. Differential gene expression with increasing β-amyloid also suggested upregulation of β-amyloid production and binding pathways in excitatory neurons in *TREM2var* heterozygotes. There was lower enrichment for pathways positively adaptive to pathology and expressed in inhibitory neurons from *CV* samples for both *TREM2var.* Exploratory bulk tissue proteomics support these observations with evidence for adaptive plasticity in response to β-amyloid pathology in *CV* tissue not found for the *TREM2var,* which showed evidence of increased β-amyloid formation and neuroplasticity changes. Together, these results highlight differences in molecular pathology between *CV* and *TREM2var* and between the *TREM2var* risk variants. They highlight mechanisms of AD risk mediated by secondary effects on astroglial and neuronal functions. Demonstration of strong epistasis between *TREM2* and *CD33* with AD supports the therapeutic potential of modulators of *CD33* inhibition or expression.

## Introduction

*TREM2* coding variants confer genetic risk for late onset AD [25]. TREM2 is expressed primarily in microglia where signalling through its adapter DAP12, which contains immunoreceptor tyrosine-based activating motif (ITAM), regulates β-amyloid clearance, lipid homeostasis and inflammatory responses [47, 69]. Expression of rare *TREM2* variant (*TREM2var*) alleles affecting the coding sequence are associated with increased AD risk [19, 51]. The *R47H* (rs75932628, minor allele frequency [MAF]=0.2%) variant confers the greatest relative AD risk (∼3.9 times that of homozygous common variant alleles [*CV*]) [16]. The *R62H* (rs143332484) variant (MAF=0.97%) has a smaller effect on disease risk (∼1.4 times homozygous *CV*) [23, 35]. Studies of human AD *post-mortem* tissue have described greater neuronal loss with both *TREM2var* compared with the *CV* [42, 48, 71]. These data suggest that the *CV* has a protective effect for AD that is reduced with risk variants. However, the complexity of the functional pathways modulated by TREM2 raises the question of whether non-cell autonomous mechanisms of pathology differ between risk genotypes.

Functional changes with *TREM2 KO* or the *R47H TREM2* allelic variant have been characterised in microglia in animal models. β-amyloid clearance is reduced in *TREM2 KO* mice [14, 43, 48, 61]. *TREM2 KO* or *R47H* human β-amyloid models both show increased neuritic plaque density and *TREM2 KO* in a humanised microglia mouse tauopathy model leads to dysregulation of neuronal stress kinase pathways and greater tau hyperphosphorylation [4]. These effects are unlikely to be mediated solely by microglial pathology given the close functional interactions between microglia, astrocytes and neurons, but the impact of expression of different risk variants in microglia on astrocytes or neurons with AD has yet to be explored.

Although evidence in mouse models have suggested epistasis between *TREM2* and *CD33*, which encodes CD33, a sialic acid binding immunoglobulin lectin (SIGLEC) [17], this has not yet been demonstrated in human AD. The rs3865444 *CD33var* is linked to reduced microglial membrane expression of CD33 protein. An AD genome wide association study has defined a small protective effect of rs3865444 *CD33* polymorphisms [20, 37]. The reduced risk of AD may in part be a consequence of reduced immunoreceptor tyrosine-based inhibitory motif (ITIM) availability on the receptor [47].

Here we have contrasted cellular pathology in human *post-mortem* brain with and without AD to test mechanisms associated with the differential genetic risks conferred by *R47H, R62H* and the *CV* with and without *CD33var*. Using immunohistochemistry (IHC) and imaging mass cytometry (IMC), we characterised neocortical β-amyloid pathology with *CV, R47H* or *R62H TREM2var*. We performed single nuclear RNA sequencing (snRNAseq) to test for differences in cell-type differential gene expression (DGE) as well as bulk proteomics in *TREM2var* and *CV* cortical tissues with greater 4G8^+^ β-amyloid immunostaining. Our work characterised *TREM2* genotype dependent differences in effects of the protective *CD33* allele on expression of microglial phenotype. We also demonstrated that the functional effects of *TREM2var* with increasing β-amyloid load in AD are associated with distinct astrocytic and neuronal response signatures. Together, this provides evidence of the mechanistic relationships between microglia, astrocytes and neurons in the genesis of AD.

## Methods

### Brain tissue

This study was carried out in accordance with the Regional Ethics Committee and Imperial College Use of Human Tissue guidelines. Cases were selected based first on neuropathological diagnosis (NDC or AD) from UK brain banks (London Neurodegeneration [King’s College London], Newcastle Brain Tissue Resource, Queen’s Square Brain Bank [UCL], Manchester Brain Bank, Oxford Brain Bank and Parkinson’s UK [Imperial College London] Brain Bank). We then excluded cases with clinical or pathological evidence for small vessel disease, stroke, cerebral amyloid angiopathy, diabetes, Lewy body pathology (TDP-43), or other neurological diseases. Where the information was available, cases were selected with a *post-mortem* delay of less than 49 h (**Table 1, Supp. Table 1**). Mid temporal gyrus (MTG) and sensorimotor (SOM) cortex from a final set of 58 cases including 16 non-diseased control (NDC) cases (Braak stage 0–II) and 42 AD cases (Braak stage III–VI) were used (total of 107 brain samples) (**Fig. 1a**, **Table 1**). Cortical samples from two regions were prepared from each brain to characterise pathology and transcript expression with both higher (mid temporal gyrus) and lower (sensorimotor) tissue densities of pathology.

**Fig. 1.**
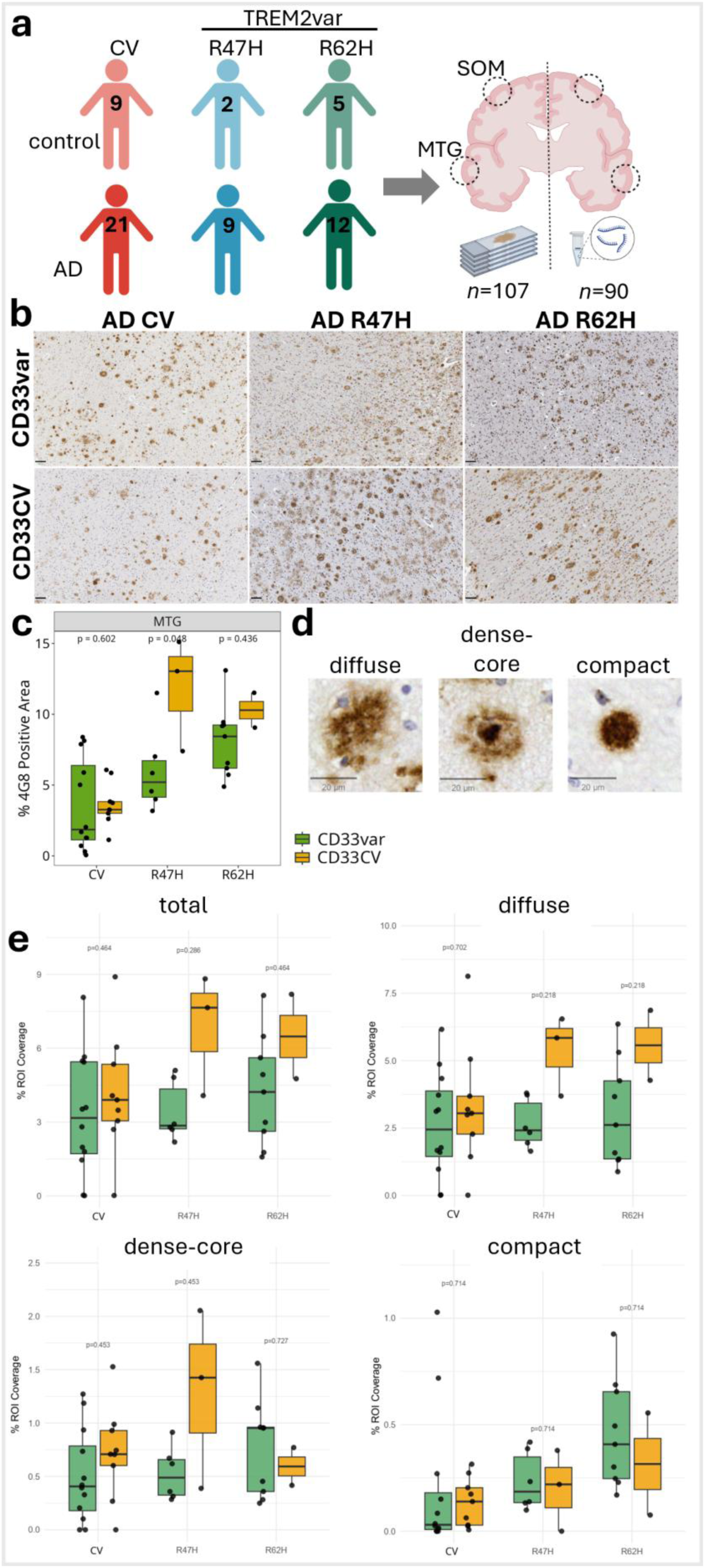
TREM2 variants lead to increases in amyloid pathology rescued by a protective CD33 polymorphism. **a** 58 post-mortem tissue blocks from donors with CV or TREM2 variant (R47H, R62H) from two brain regions (mid-temporal gyrus [MTG] and somatosensory cortex [SOM]) were studied. Formalin fixed, paraffin embedded sections (FFPE) were used for histopathology and paired frozen tissue from the homologous regions in the contralateral hemisphere of the same brain was used for transcriptomics in each case. **b** Representative images of 4G8 β-amyloid staining of human post-mortem tissue (scale bars = 100 µm) stratified by TREM2 and CD33 genotypes. **c** Increased β-amyloid with TREM2var in MTG of AD cases, particularly R47H, was partially rescued by the CD33 protective variant (CD33var) compared to the CD33 CV group. **d** Plaques were characterised by morphology as diffuse, dense core or compact **e** Plaque coverage analysis for total, diffuse and dense-core plaques, stratified by TREM2 and CD33 genotype, showed a trend towards a reduced total plaque coverage and area covered by diffuse and dense-core plaques with CD33var in R47H tissue.

**Table 1.**
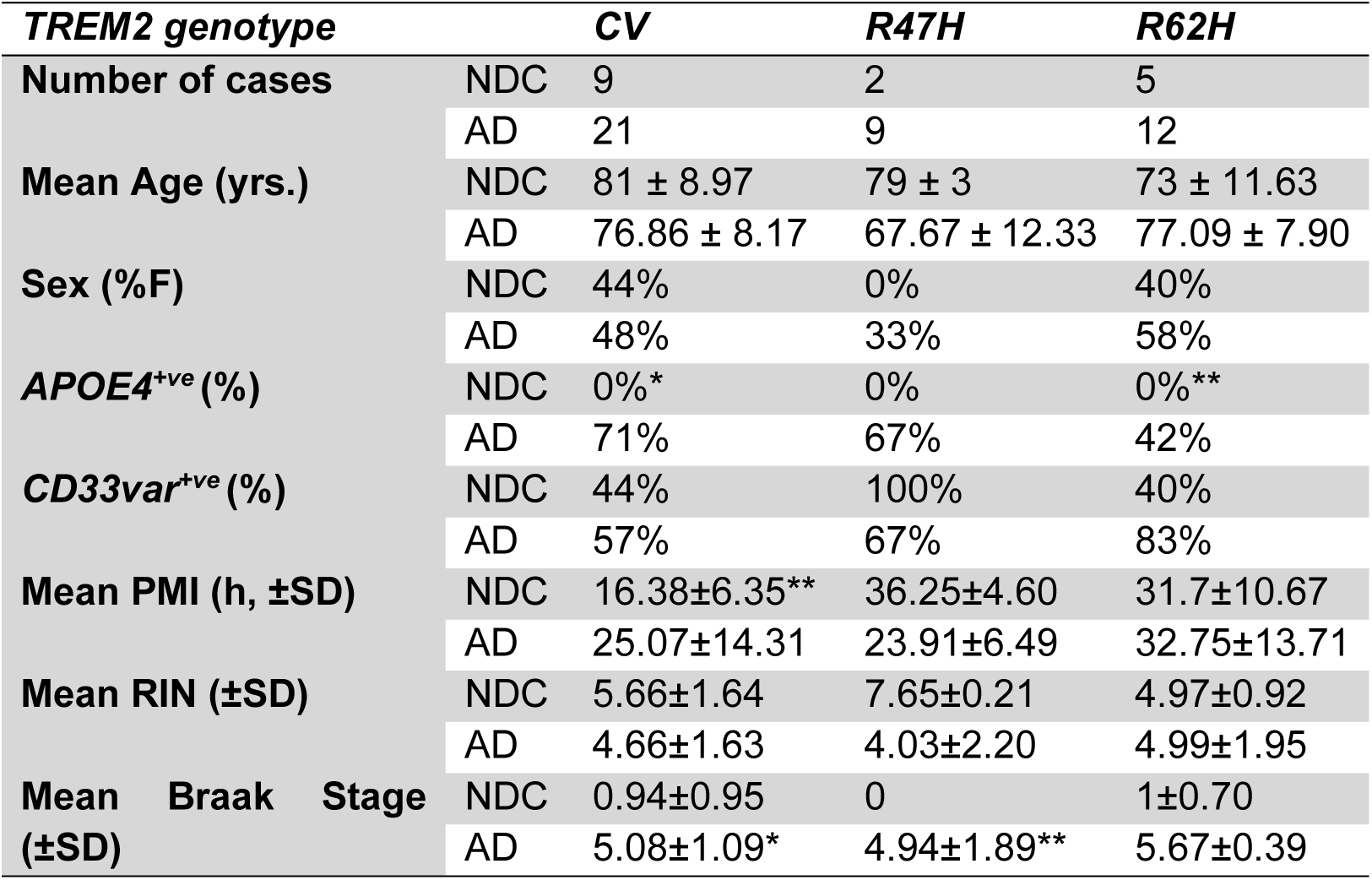
Demographic characteristics and neuropathological findings across genotypes and disease groups. Post-mortem interval (PMI). RNA integrity value (RIN). Mean ± SD. *unknown for 3 cases, **unknown for 1 case.

### Genotyping

DNA was extracted from frozen brain tissue using the Qiagen DNeasy Blood and Tissue Kit. All samples were genotyped on the UK Biobank Axiom Array with a subset confirmed on the Illumina NeuroBooster Array (NBA). Standard quality control procedures were performed in PLINK (v1.9) and R (4.0.5, 2021-03-31) at the individual sample level and then the single nucleotide polymorphism (SNP) level. Relevant SNPs (*CD33*, *APOE*, *TREM2*) were extracted to determine genotype for each locus for each sample.

### Immunohistochemistry (IHC)

IHC was performed on FFPE sections (n=107) from the MTG and SOM of each brain studied and paired with material from the cryopreserved contralateral hemisphere used for nuclear preparations for snRNA sequencing. Standard immunostaining procedures as recommended by the manufacturer were followed a Super Sensitive Polymer-HRP (Biogenex) kit. Briefly, after dewaxing and rehydration of slides, endogenous peroxidase activity was blocked with 0.3% H_2_O_2_, followed by antigen retrieval using a steamer and citrate buffer (pH 6). Primary antibody for β-amyloid (4G8; Biolegend), which immunolabels amyloid plaques, intracellular amyloid, and APP and its cleaved products [21], was incubated overnight at 4 °C. Species-specific Super Sensitive kits and DAB were used for antibody visualisation. Tissue was counter-stained by incubation in Mayer’s haematoxylin (TCS Biosciences) for 2 min, followed by dehydration, clearing and mounting.

### Image Analysis

Digital images were generated from IHC stained slides scanned using a Leica Aperio AT2 Brightfield Scanner (Leica Biosystems). Images were analysed using Halo software (Indica Labs) after optimisation of Indica Labs macros for each antibody (β-Amyloid = Area Quantification v1). Data from both cortical regions in each brain were combined. Data was analysed using R package with Wilcoxon signed-rank test and confirmed with a linear regression model incorporating APOE genotype as covariate.

### Microglial Activation Morphology analysis

IHC images from Iba1 (WAKO, 019-19741) staining were analysed using the Microglial Activation Module in Halo software. The module was optimized to detect nuclei from hematoxylin staining and microglial somata and processes from Iba1 staining by defining signal thresholds. An additional threshold (1.9x) for the ratio of soma area to process area was manually defined to classify microglia as having activated or non-activated morphology. The percentage of activated microglial cells was calculated as the number of activated microglia divided by the total number of activated and non-activated microglia. Data were analysed in R using the Wilcoxon signed-rank test.

### Plaque subtype analysis

Plaque density was assessed as an additional analysis specifically in the subset of cases that passed snRNAseq QC for inclusion in the transcriptomic analysis. Images of whole slides immunostained with 4G8 were uploaded to Biodock [71], where β-amyloid deposits were characterised into four groups based on morphology: diffuse, dense cored, compact, and cerebral amyloid angiopathy. 2×2mm tiles were labelled using the semi-automated AI prelabelling function, followed by manual correction. 152 fully labelled tiles were used to train a deep learning model within Biodock. After several series of labelling and training and quality accuracy checks, the final model was applied to the full cohort and plaques above a confidence threshold of 0.53 were extracted for analysis. Plaques within a defined rectangular region of interest within the grey matter were taken forward to analysis.

Data analysis was performed in R Studio. Plaques with a diameter less than 10µm were excluded. Densities (plaques per square mm) for each plaque type were calculated as the total plaque number per unit area (mm^2^). Percentage coverage by plaques was calculated as the total plaque area, within each subtype and overall, divided by the ROI area. Plaque densities and coverage were compared between groups using Wilcoxon tests, with p values adjusted for multiple comparisons using Benjamini-Hochberg correction. Plaque size comparison was performed on all recognised plaques, without a minimum size threshold. Within each *TREM2* genotype, inverse cumulative plaque size distributions between *CD33* variants were compared using Kolmogorov-Smirnov tests.

### Imaging Mass Cytometry (IMC)

FFPE 5-10μm sections from MTG and SOM were immunostained using lanthanide tagged antibodies (**Table 2**) before ablation. The slides underwent routine dewaxing and rehydration before undergoing antigen retrieval, in a pH8 Ethylenediaminetetraacetic acid (EDTA) buffer. The slides were then treated with a 10% normal horse serum (Vector Laboratories) blocking solution before incubation with an antibody cocktail (Table 3) at 4⁰C overnight. The slides were washed in 0.02% Triton X-100 (Sigma-Aldrich) before incubation with the Iridium-intercalator (Standard BioTools) then washed and air-dried. All antibody conjugation was performed using the Maxpar X8 protocol (Standard BioTools).

**Table 2:** Antibodies used for imaging mass cytometry.

| Antibody | Manufacturer | Ln-Isotope | Concentration |
| --- | --- | --- | --- |
| **GFAP** | Fluidigm, 3143030D | 143Nd | 1:500 |
| **β-Amyloid (4G8)** | Biolegend, 800711 | 144Nd | 1:500 |
| **S100β** | Fluidigm, 3146021D | 146Nd | 1:2000 |
| **MAP2** | Fluidigm, 3148023D | 148Nd | 1:400 |
| **CD68** | Fluidigm, 3159035D | 159Tb | 1:200 |
| **PVALB** | Novus, NB120-11427 | 163Dy | 1:1000 |
| **Iba1** | WAKO, 019-19741 | 169Tm | 1:500 |
| **HLA-DR** | Fluidigm, 3174025D | 174Yb | 1:200 |

IMC was performed using a Hyperion Imaging Systems First Generation (Standard BioTools) coupled to a CyTOF First Generation mass cytometer (Standard BioTools). The instrument was first tuned using the manufacturer’s 3-Element Full Coverage Tuning Slide before the slides were loaded into the device. Four 500×500μm^2^ regions of interest within the grey matter were selected and ablated using a laser operating at a frequency of 200Hz with 1μm resolution. Data were stored as .mcd files and processed using the steinbock pipeline (Bodenmiller Group). Hot pixels were removed, and images were exported as TIFF files. Pixel classification was performed using the Ilastik module to generate nuclear masks based on DNA staining and cytoplasmic masks based on combined signals from GFAP, S100B, MAP2, PVALB, CD68, Iba1, and HLA-DR (Table 2). Nuclear and cytoplasmic masks were subsequently combined using the CellProfiler module to generate single-cell masks. For each cell mask, the aggregated median intensity of each channel was quantified. The resulting single-cell data were imported into imcRtools (Bodenmiller Group) for visualization and downstream analysis.

### Quantification of Astrocytes Using IMC

IMC images generated using metal-conjugated antibodies against GFAP and S100β (Table 2) were used to determine the density and proportion of GFAP⁺ cells among GFAP⁺S100β⁺ astrocytes in MTG and SOM regions from AD and NDC cases. Thresholds for GFAP and S100β signals were manually set in imcRtools for each case in a genotype-blinded manner. Cells with signal intensities above these thresholds were classified as GFAP⁺ and/or S100β⁺ astrocytes. Astrocyte counts from four 500 × 500 μm² ROIs per case were summed to calculate cell density per mm². A Wilcoxon signed-rank test was used for testing the significance of differences between the *CV* and *TREM2var* groups.

### Single nucleus isolation for single nuclear RNA sequencing (snRNAseq)

Single nucleus isolation was conducted on 90 tissue blocks from CV (48 total: *n*=18, NDC [9 MTG, 9 SOM]; *n*=21, AD [19 MTG, 11 SOM]) or either the *R47H* (17 total: *n*=2 [2 MTG], NDC; *n*=15, AD [8 MTG, 7 SOM]) or *R62H* (25 total: *n*=7 [5 MTG, 2 SOM], NDC; *n*=18, AD [10 MTG, 8 SOM]) *TREM2var* (**Supp. Table 4**). Homologous fresh frozen brain tissue blocks from the MTG and SOM were cryosectioned at 80µm and 200mg of grey matter was collected in RNAse-free Eppendorf tube, as previously described [52]. Nuclei were isolated as previously described [52] using a protocol based on [30]. All steps were carried out on ice or at 4°C. Tissue was homogenised in homogenisation buffer (1% Triton X-100, 0.4 U/μl RNAseIn + 0.2 U/μl SUPERaseIn, 1µl 1mg/ml DAPI) using a 2ml glass douncer. Homogenate was centrifuged at 4°C for 8 min, 500g and supernatant removed. Homogenate was resuspended in homogenisation buffer and filtered through a 70µm filter followed by density gradient centrifugation at 13,000g for 40 min. Supernatant was removed and nuclei were washed and filtered in PBS buffer (PBS + 0.5mg/ml BSA + 0.4 U/μl RNAseIn + 0.2 U/μl SUPERaseIn). Nuclei were pelleted, washed twice in PBS buffer and resuspended in 1ml PBS buffer. 100µl of nuclei solution was set aside on ice for single nuclear processing.

### Single nucleus processing and sequencing

Isolated nuclei stained with Acridine Orange dye were counted on a LUNA-FL Dual Fluorescence Cell Counter (Logos Biosystems, L20001). Sufficient nuclei for recovery of 7000 cells were used for 10x Genomics Chromium Single Cell 3ʹ processing and library generation. All steps were conducted according to the 10x Genomics Chromium Single Cell 3’ Reagent Kits v3 User Guide, with 8 cycles of cDNA amplification until fragmentation, where 25ng of amplified cDNA per sample was taken through for fragmentation. The final index PCR was conducted at 14 cycles. cDNA and library prep concentration were measured using Qubit dsDNA HS Assay Kit (ThermoFisher, Q32851) and DNA and library preparations were assessed using the Bioanalyzer High-Sensitivity DNA Kit (Agilent, 5067-4627). Pooled samples at equimolar concentrations were sequenced on an Illumina HiSeq 4000 according to the standard 10X Genomics protocol.

The full snRNAseq data will be made available for download from the Gene Expression Omnibus (GEO) database (https://www.ncbi.nlm.nih.gov/geo/) upon publication.

### Pre-processing and quality-control of snRNA sequencing data

Alignment and demultiplexing of raw sequencing data was performed using 10X Genomics Cell Ranger (v7.1.0), with a pre-mRNA GRCh38 genome reference including both introns and exons. Ambient RNA contamination was removed from the raw feature-barcode matrices using CellBender (v0.3.0) (remove-background) with default parameters. The CellBender output filtered feature-barcode matrices were used for downstream primary analyses using our scFlow pipeline (v0.7.5) [27]. Cells were filtered for ≥500 and ≤20000 total counts and ≥400 and ≤8000 total expressive features, where expressivity was defined as a minimum of 1 count in at least 3 cells. The maximum proportion of counts mapping to mitochondrial genes was set to 10%. Doublets were identified using the DoubletFinder algorithm, with a doublets-per-thousand-cells increment of 8 cells (recommended by 10X Genomics), a pK value of 0.005, and embeddings were generated using the first ten principal components calculated from the top 2000 most highly variable genes (HVGs) [38].

### Integration, clustering, and visualization of data

The linked inference of genomic experimental relationships (LIGER) package (rliger v2.2.1.9001) was used to calculate integrative factors across samples [65]. LIGER parameters used included: k: 40, lambda: 5.0, max_iters: 30, knn_k: 20, min_cells: 2, quantiles: 50, num_genes: 2000, center: false. Two-dimensional embeddings of the LIGER integrated factors were calculated using the uniform-manifold approximation and projection (UMAP) algorithm (R package uwot v0.2.4.9000) with the following parameters: pca_dims: 50, n_neighbours: 35, init: spectral, metric: euclidean, n_epochs: 200, learning_rate: 1, min_dist: 0.4, spread: 0.85, set_op_mix_ratio: 1, local connectivity: 1, repulsion_strength: 1, negative_sample_rate: 5, fast_sgd: false [38]. The Leiden community detection algorithm was used to detect clusters of cells from the 2D UMAP (LIGER) embeddings; a resolution parameter of 0.0001 and a k value of 50 was used [57].

### Assigning cell type labels to snRNAseq cells

Automated cell-typing was performed essentially as previously described using the Expression Weighted Celltype Enrichment (EWCE v1.19.0) algorithm in scFlow against a previously generated cell-type data reference from the Allan Human Brain Atlas [27, 51] (**Supp. Table 5**). The top five marker genes for each automatically annotated cell-type were determined using Monocle 3 and validated against canonical cell-type markers [58].

### Differential gene expression analysis

We used a pseudocell-based strategy combined with linear modelling to identify differentially expressed genes from snRNA-seq data while mitigating sparsity and technical variability [15]. Within each donor and cell type, raw UMI counts from approximately 30 randomly selected nuclei were aggregated to form pseudocells. For donor–cell type combinations pseudocells with fewer than 10 nuclei were excluded.

Differential expression analyses were performed using the **limma-trend** framework with robust empirical Bayes moderation. Pseudocell count matrices were normalised to counts per million (CPM) to account for differences in library size, followed by quantile normalisation to equalise expression distributions across pseudocells. Gene expression was modelled jointly across all samples using a linear regression model that included continuous pathology measures (pct4G8PositiveArea) and categorical *TREM2var* status (*CV, R47H, R62H*) within a single design matrix. Sex, Brain Region, *APOE4* status, *CD33* genotype status, scaled pseudocell size, log2-transformed pseudocell mitochondrial proportion, and log2-transformed pseudocell UMI counts were included as covariates. Donor identity was included as a random effect to account for repeated measures across pseudocells. Differentially expressed genes were identified based on absolute log2 fold-change (>0.25) and false discovery rate (FDR)–adjusted p values (≤0.05) derived from moderated t-statistics.

### Impacted pathway analysis

Impacted pathway analysis (IPA) was performed using enrichR packages in scFlow [9, 35]. Statistically significant differentially expressed genes were used against the Gene Ontology (GO) databases. Results were ordered by the enrichment score, defined as the proportion of genes overlapping with a pathway relative to the expected overlap. Pathways were selected based on the removal of pathways with FDR<0.05. Pathways with gene overlapped fewer than 3 were excluded.

### Proteomics

OCT from fresh-frozen tissue was removed via sequential washing in ethanol and ammonium bicarbonate based on Valdés *et al.* [61]. Frozen tissue was sectioned into 10 2×80µm sections into a 2mL Protein LoBind Eppendorf with 1mL ice-cold 70% (V/V) ethanol and rotated end-over-end at 4°C for 2 min. This was repeated for a total of 5 washes, centrifuging at 10,000xg, 4°C for 2 min and discarding the supernatant between washes. Similarly, 4 washes with 1mL water and 3 washes with 50mM ammonium bicarbonate were conducted and the sections allowed to air dry for 5 min. Protein was extracted by the addition of 50µL 8M urea in 100mM ammonium bicarbonate and 1mM phenylmethylsulfonyl (PMSF) with probe sonication on ice with 10×1s pulses at 30% amplitude. The protein was recovered from the supernatant by centrifuging for 10 min at 10,000xg and freezing at -80°C. 20μg of protein was reduced, alkylated and digested using an in-solution digestion approach automated on an Andrews+ pipetting robot with a 96-PCR Plate Peltier+ (Waters). Briefly, at 4°C, 20μg of protein was combined with water and a pre-mixed digestion solution, resulting in 0.2μg trypsin (Sequencing Grade Modified Trypsin, Promega), 100mM ammonium bicarbonate, 40mM chloroacetamide and 10mM Tris(2-carboxyethyl)phosphene and then heated to 37°C for 16 hours before being chilled to 4°C and acidified to 0.5% (V/V) trifluoro-acetic acid (TFA). Peptides were desalted in a semi-automated manner using an Andrews+ pipetting robot in combination with a Vacuum+ (Waters) by solid-phase extraction on an Oasis HLB μElution Plate (Waters). Briefly, the plate was pre-equilibrated by washing three times with 100μL 0.1% (V/V) TFA. All samples were added to the plate and washed a further 3 times with 100μL 0.1% (V/V) TFA before being eluted with 50μL followed by 100μL 60% acetonitrile into 96-well QuanRecovery plates (Waters) and dried at 60°C using a Speedyvac (Eppendorf). All volumes were drawn through the plate by negative pressure. All samples were reconstituted into 200μL 0.1% formic acid (FA) by sonicating for 10 min in a water bath. A pooled sample was generated from 20μL of all samples and used as a study reference (SR) and for spectral-library generation. LC-MS analysis was performed on an M-Class (Waters) hyphenated to a 7600 ZenoTOF (Sciex) with peptide separation across a Kinetex 2.6µm XB-C18 150×0.3mm column (Phenomenex). 3µL of sample was injected, with samples interspaced with a pooled study reference every 5 samples. The LC ran from 3% buffer B (acetonitrile, 0.1% FA), 97% A (water, 0.1% FA) to 30% B over 20 min at 5µL/min. Columns were washed with 80% buffer B and equilibrated to 3% buffer B for 4 column volumes between injections. Masses were acquired between 400 and 1500mz (MS1) and 140 to 1800mz (MS2) following CID fragmentation. MS2 accumulation time was 0.013s with Zeno pulsing and dynamic collision energy enabled. Precursor ion selection was performed in data-independent acquisition across 85 variable windows between 399.5mz and 903.5mz. LC-MS instrumentation was controlled in Sciex OS 3.0 (Sciex).

Data was searched in DIANN 2.2 with precision QuantUMS quantitation [29] and DIANN set to optimise MS1, MS2 and scan window thresholds. Peptides and proteins were filtered using default FDR settings (1% global FDR, 5% run-specific FDR for proteins) and match-between runs was enabled. Data was searched against a spectral library, generated from fractionated pooled sample reference containing 77,182 precursors from 5,243 proteins. To generate the spectral library, samples from multiple rounds of fractionation were combined and searched. Pooled sample from this study was fractionated into 11 using High pH Fractionation Kit (Pierce) as per the manufacturer’s instructions. Fractions were injected twice using data dependant acquisition. In the first injection up to 60 precursors were selected for fragmentation between 500 and 655mz. In the second injection, precursors were selected between 455 and 905mz. MSMS accumulation time was 0.01s and masses were recorded between 140 and 1800mz with dynamic collision energy. Additional offline high pH reversed phase fractionation was performed from 100µg of pooled peptides from this study, and pooled cytosolic fraction from synaptosome enrichment [18], separated over a 45 min gradient of 4% Buffer B (4mM ammonium bicarbonate 90% (V/V) acetonitrile) to 72% Buffer A (4mM ammonium bicarbonate, 2% (V/V) acetonitrile) with a bioZen 3µm NX-C18 150×2.1mm (Phenomenex) column with fractions collected every minute. Finally, 100µg of proteins from the cytosolic fraction pooled sample was separated on a 4-12% Bis-Tris 12-WedgeWell Midi Gel (NuPage, Thermofisher), cut into 15 bands and subjected to Shevchenko in-gel digestion [49]. Peptides were analysed by DDA, with separation across a Kinetex 2.6µm XB-C18 150×0.3mm column (Phenomenex), with a gradient spanning 3% Buffer B (acetonitrile, 0.1% (V/V) formic acid) to 70% Buffer A (Water, 0.1% (V/V) formic acid) over 45 min. .wiff files were converted to .mzML and centroided using MSConvert (Proteowizard) and searched using Fragpipe version 19.0 with MSFragger version 3.6 and default settings for Basic-Search with spectral library generation enabled. Data was searched against the canonical human proteome downloaded from Swissprot (Uniprot.org) on 07/12/2022 and supplemented with a decoy library automatically in Fragpipe. The mass spectrometry proteomics data have been deposited to the ProteomeXchange Consortium via the PRIDE [45] partner repository with the dataset identifier PXD074536.

## Results

### Increased β-amyloid plaque accumulation in TREM2var carriers is modified by CD33

We performed IHC and snRNAseq on paired hemispheric formalin-fixed and frozen blocks from the same brains for these studies (**Fig. 1a, Supp. Table 2-4**). Immunohistology of *TREM2 R47H* and *R62H* sections showed increased 4G8^+^ β-amyloid staining in cortical sections from AD donors carrying *TREM2* variants compared to those carrying *CV* with no *APOE4* effect on amyloid pathology (**Fig. 1b, Supp. Fig. 1a, b**).

We tested for epistasis between *CD33* and *TREM2* [17] by sample stratification for both genotypes. We first tested for differences in 4G8^+^ immunostaining as a measure of total β-amyloid plaques in in the mid-temporal gyrus (MTG) with AD (**Fig. 1c**). The 4G8^+^ area did not differ significantly by *CD33* genotype for *CV*, but a 2.5-fold lower 4G8^+^ immunostaining area was found in *R47H* carriers heterozygotic for the protective *CD33var* (**Fig. 1c**). We conclude that *CD33* polymorphisms can partially compensate for functional impairments in β-amyloid clearance associated with *TREM2var*. We next conducted an exploratory analysis of differences in total 4G8^+^ plaque densities and plaque subtype densities (diffuse, compact and dense core plaques; **Fig. 1d**) among *TREM2* genotypes. There were consistent trends for higher total and compact plaque density for the *TREM2va*r (**Supp. Fig. 1c**). We next observed that *CD33var* trended towards rescuing plaque coverage to a similar extent (2.7-fold lower total plaque coverage in *R47H CD33var* vs *R47H CD33 CV*) as in the total 4G8^+^ area particularly for diffuse and dense-core plaque subtypes (**Fig. 1e**). This was further supported by a reduction in overall plaque size across *TREM2* genotypes with *CD33var*, most apparent in *R47H* cases (**Supp. Fig. 1d, e**).

In summary, we found increased β-amyloid load alongside trends for increases in total plaque densities with *TREM2var,* suggesting either decreased β-amyloid clearance or increased production, and modulated by *CD33var*. The increased β-amyloid load was associated with a greater proportion of compact plaques in the *TREM2var* heterozygotes.

### TREM2 genotype-dependent differences in both microglial and astroglial activation with AD

We hypothesised that microglia with heterozygous *TREM2var* expression show different activation responses to β-amyloid. To test for this, we assessed genotype dependent differences in pro-inflammatory activated microglia between MTG and sensorimotor cortex (SOM) tissue, which expresses pathology relatively later in AD progression. There were consistent trends for an increase in activated microglial morphologies with AD in both *CV* MTG and SOM (**Fig. 2a, b**). However, while there was a greater mean proportion of microglia with activated morphology with the *R62H* in the MTG, there was a trend to reduced activation with *TREM2var* in the SOM (**Fig. 2a, b**). We did not find evidence for effects of the protective *CD33var*. The density of total (GFAP^+^ or S100β^+^) and proportion of potentially reactive GFAP^+^ astrocytes showed similar trends to those of the microglia. There were greater total numbers of astrocytes with AD for *CV* (p<0.05) with a trend towards even higher numbers with the *R47H* genotype. However, the mean GFAP^+^ astrocyte proportion of total astrocytes was higher in *R62H* than for *CV* or *R47H* (**Fig. 2c, d**). Fewer SOM samples with *CD33 CV* precluded meaningful exploration of regional differences related to *CD33*.

**Fig. 2.**
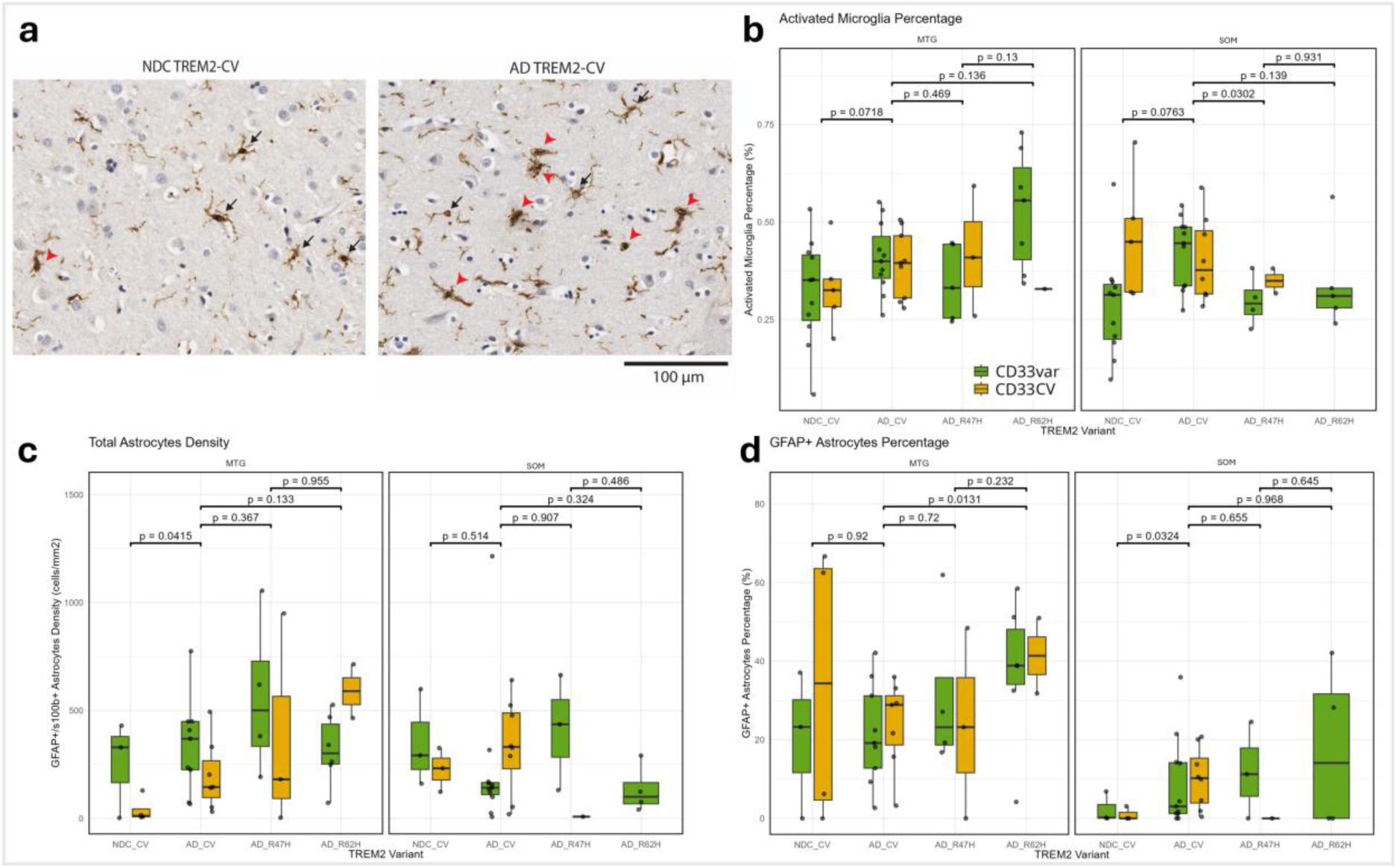
Relative microglial and astrocyte densities in non-diseased and AD tissue vary by TREM2 genotype. **a** Immunohistochemistry showed an increase in microglia with an activated morphology (red arrow heads) compared to ramified microglia (black arrows) in AD CV cases vs non-disease controls (NDC). **b** The proportion of activated microglia was consistent across genotypes in MTG but not SOM. **c** Total astrocyte density increased in AD in MTG as assessed by IMC. **d** The proportion of activated GFAP+ astrocytes was increased in R62H MTG cases. Here, both CD33 genotypes were combined to assess the effect of TREM2var status.

We thus found evidence for greater pro-inflammatory microglia activation with AD relative to non-diseased *CV* tissue with the *R62H TREM2* genotypes in the MTG but not at the earlier stage of pathology reflected in the SOM. IHC also provided evidence for astrogliosis with the variant genotypes in both the MTG and SOM, highlighting a possible role for astrocytes in expression of *TREM2var* pathology.

### Transcriptomic evidence for epistasis between TREM2 and CD33

We further explored the apparent epistasis between *TREM2* and *CD33* suggested by IHC using snRNAseq using a joint linear regression model. We assessed *CD33*-dependent differential expression in microglia with greater β-amyloid deposition by comparing subgroups carrying *CD33* common allele only and with those heterozygous for the *CD33var* within each *TREM2* genotype. We then compared the CD33-dependent genes in each *TREM2var* group relative to the *CV* group (**Supp. Table 34**). We found more genes whose expression was modulated by the *CD33* genotype difference in the *R62H* than in the *CV* or the smaller *R47H* sample set with a trend *R62H>CV*>*R47H* (numbers of significantly differentially expressed genes: 54 *R62H*, 16 *CV* and 0 *R47H*) (**Fig. 3a, b**). CD33-dependent genes in *R62H* included genes associated with microglial activation (*PTPRG, SSH2, PLTP, HM13*) [38]. Whilst R47H microglia did not have any differentially expressed genes which survived multiple comparison testing, there were a number of differentially expressed genes significant at the nominal level, including a number of those involved in microglia activation (*GLUL, LMO4, SYT17, LILRB1*) [38].

**Fig. 3.**
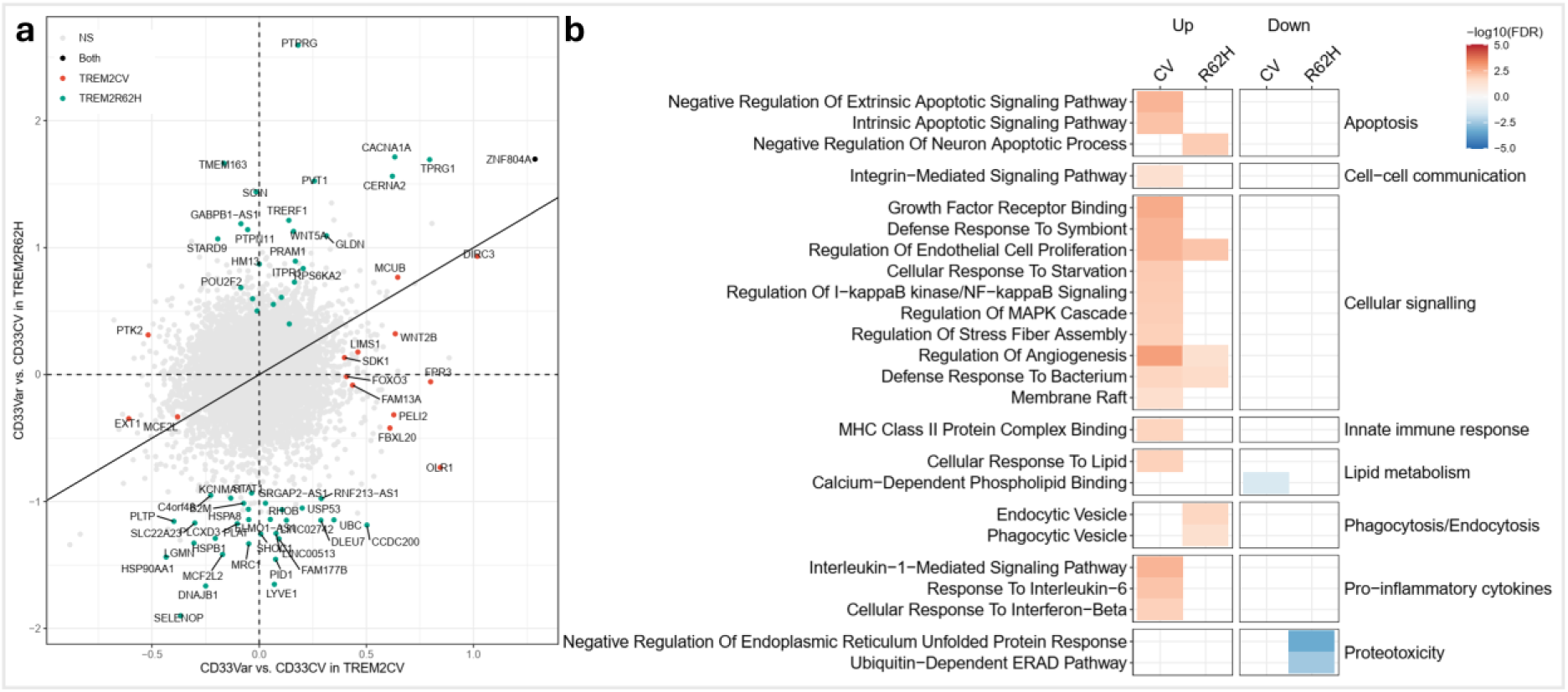
TREM2 genotype dependent effects of the protective CD33 polymorphism on microglial responses to increasing amyloid load. **a** DEG analysis of CD33-TREM2 interaction identified CD33-dependent genes in CV and R62H but not R47H cases. Axes are Log_2_FC between CD33 variants. Pathology load was corrected for to assess microglial responses across genotypes where pathology load differs. **b** Reduced activation responses of microglia with TREM2var to β-amyloid. Microglial activation was reduced for R62H and was too low for identification of pathways with R47H.

We performed pathway analyses for the (fully corrected) significant DEG for *CD33* allelic differences within each *TREM2* genotype (**Supp. Fig. 2**). We then clustered the differentially expressed genes based on associations to previously described *TREM2* functions in microglia. Only one of the enriched pathways was similar across *R62H* and *CV* genotypes, ‘transcriptional regulation’. This observation, combined with the increase in numbers of genes with expression modulated by the *CD33* genotype in *R62H* suggests a substantial functional impact on residual*TREM2*-mediated signalling. However, most pathways were represented only for one genotype. For example, *R62H* uniquely showed *CD33* modulation for ‘cell signalling’ (*PTRG, PRAM1*), ‘microglial activation’ (*TMEM163*) and ‘phagocytic and endocytic’ pathways. These included genes associated with disease-associated microglia (*B2M*) [27] and plaque-induced genes (*LGMN*) [70].

These results provide additional insights into differences between *R47H* and *R62H* associated *TREM2* functional impairments. They highlight the extent to which the protective *CD33* genotype may normalise microglial transcriptomic signatures.

### Differential transcriptomic responses to increasing β-amyloid in R47H and R62H microglia

Having confirmed a *CD33var*-dependent effect on the expression of the functional phenotypes associated with *TREM2var*, we then directly characterised the relative β-amyloid density dependent differential gene expression for microglia using a mixed effect model controlling for *CD33* and *APOE4* genotypes. We found decreased numbers of DEG with *TREM2var* (*CV* (158 DEG) > *R62H* (37 DEG) > *R47H* (4 DEG)) consistent with an attenuated transcriptional response to β-amyloid of microglia expressing the variant proteins, most markedly with *R47H* (**Supp. Fig. 3a, b; Supp. Table 18**). We next quantified response directionality using the ratio of up- to down-regulated genes expressed (R_g_). This ratio was at unity for *R47H*, but increased for *R62H* and reduced response for *CV* samples (*R62H* (R_g_ = 2.6) > *R47H* (R_g_ = 1) > *CV* (R_g_ = 0.4)). Together, these data support differential variant-specific microglia activation responses to β-amyloid.

We next explored the nature of pathways significantly differentially enriched across genotypes (**Fig. 3c; Supp. Table 31**). Genes related to pathways upregulated for *CV* samples included transcripts involved in the pro-inflammatory response such as *STAT1* involved in the ‘cellular response to interferon β’, *IL-1RAP* and *IL-1β* involved in the ‘interleukin-1-mediated signalling pathway’ and *HSPA8* involved in ‘MHC Class II Protein Complex Binding’.

The *R62H* heterozygote samples showed increased expression of *HLA-A* and *SRGAP3* transcripts involved in ‘phagocytotic vesicle’, and *APOE* and *PICALM* involved in extracellular ‘β-amyloid binding’ [29, 40]. By contrast, *R62H* heterozygotes most prominently showed a reduction in *NCK2* which is involved in the ‘negative regulation of endoplasmic reticulum unfolded protein response’ and previously shown as a response to β-amyloid oligomers [38] (**Fig. 3c**). With little transcriptional response to increasing β-amyloid detected, we did not find any pathways enriched significantly for *R47H* heterozygotes (**Fig. 3c**). These suggest reduced activation responses to β-amyloid for *R47H* and altered immune responses in *R62H* relative to CV. Primary mechanisms but which the two variants increase disease risk may therefore be different.

### Increased astrocyte transcriptional responses in TREM2var heterozygotes relative to CV

*TREM2* is primarily expressed in microglia but we observed trends to increased proportions of GFAP^+^ activated astrocytes with AD by IHC (**Fig. 2**). We therefore tested for differences in *TREM2* genotype-associated differential gene expression in astrocytes. We found that astrocytes showed genotype-dependent differences in relative activation signatures with increasing β-amyloid. Numbers of DEG were highest for the *R62H* ((*R62H* (396 DEG) > *CV* (392 DEG) > *R47H* (159 DEG)) (**Supp. Fig. 3a, c; Supp. Table 6**). However, astrocytes in *CV* (R_g_ = 1.7) and *R47H* (R_g_ = 1.6) tissues had a greater proportion of upregulated genes relative to those in *R62H* (R_g_ = 0.9) tissue.

Related pathways were increased in expression in astrocytes with greater tissue β-amyloid across the *TREM2* genotypes (**Fig. 4; Supp. Table 22**). For example, *CV*, *R47H* and *R62H* astrocytes increased expression of *APOE*, *ITM2C, CLU* and *CST3,* which contribute to ‘β-amyloid binding’ and *R47H* astrocytes strongly upregulated transcripts (*GJA1, CACNA1B)* associated with the functionally related ‘response to β-amyloid’ and related pathways associated with a β-amyloid responsive activated phenotype [11, 26]. For *R62H*, transcripts such as *NFKBIA* contributed to the upregulation of ‘regulation of NIK/NF-kappaB signalling’, a regulator of the pro-inflammatory astrocytic signature that may facilitate β-amyloid clearance [24].

**Fig. 4.**
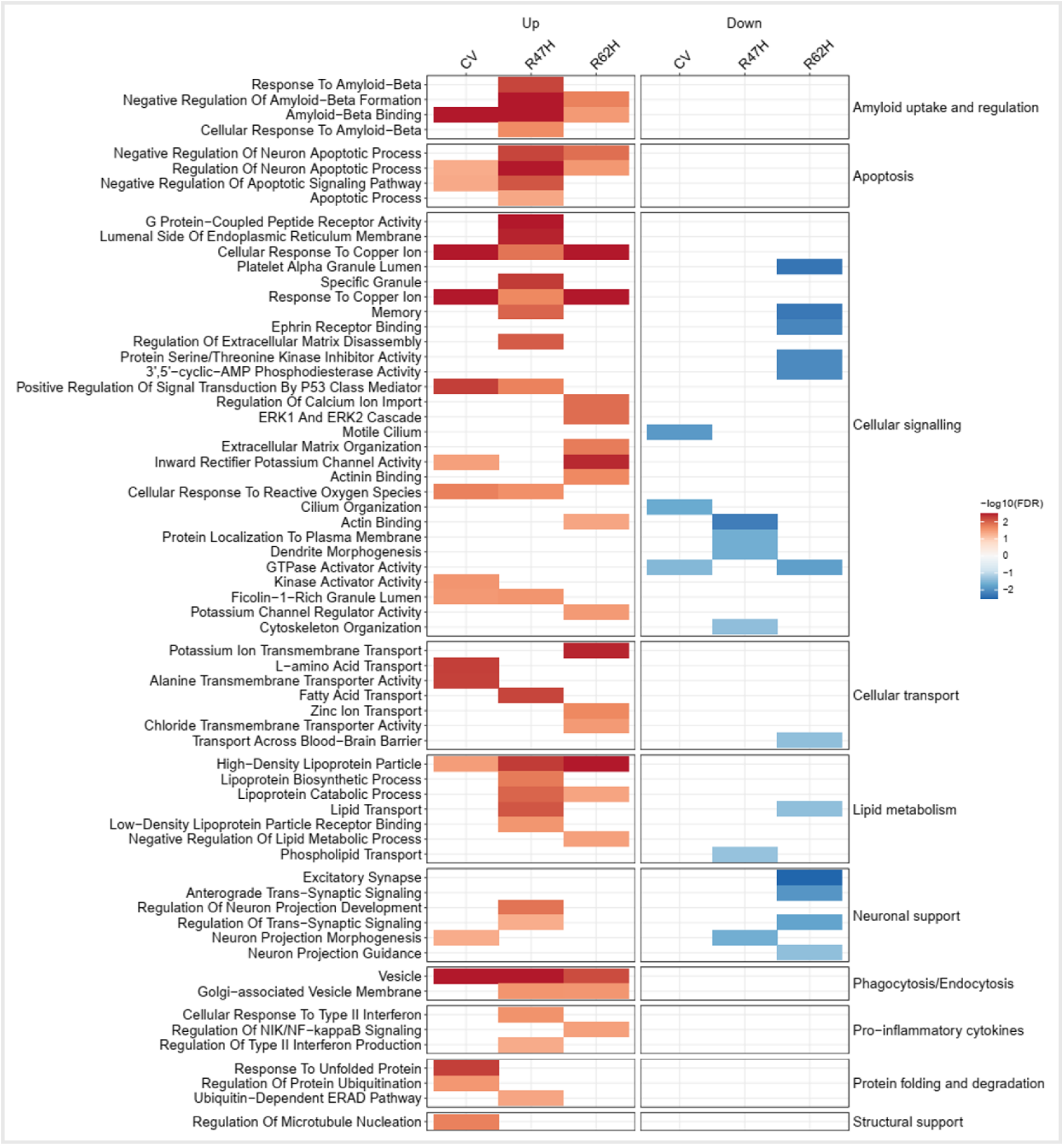
Differential gene expression of astrocytes with different TREM2 variants. Astrocytes in all genotypes showed an upregulation of a range of pathways but were enriched for pathways in ‘response to unfolded protein’ in CV, and ‘response to IFNβ’ in R47H. R62H astrocytes down-regulated pathways associated with neuronal support.

However, there also were differences. For example, *CV* astrocytes, unlike those in *TREM2var* tissues, upregulated transcripts associated with ‘response to unfolded protein’ (*HSPB8, HSPA1A*) and ‘regulation of protein ubiquitination’ (*UBB*). Astrocytes play a critical role in supporting adaptive responses in the context of neurodegeneration [13]. *CV* astrocytes upregulated transcripts for guidance molecules (*DCLK1*, *S100A6*, *ADGRB1*) involved in ‘neuron projection morphogenesis’ and those (*HSPA1A/B, PDE4DIP*) involved in the ‘regulation of microtubule nucleation’. Conversely, ‘neuron projection morphogenesis’ was downregulated in *R47H* astrocytes as were ‘actin binding’ and ‘cytoskeleton organisation’. *R62H* astrocytes down-regulated transcripts associated with the ‘excitatory synapse’ (*GRIA2, SHISA6*) and ‘anterograde trans-synaptic signalling’ (*GRM3, GABRAB1, GLRB, NRXN1, SLC1A3).* Together, these transcriptional signatures suggest potential contributions to β-amyloid clearance common to all three *TREM2 genotypes* and genotype-dependent differences in gene expression associated with adaptive plasticity of astrocytes, which showed lower expression in the *TREM2var*. They highlight a major impact of astrocyte responses on expression of the cellular pathology of *TREM2var* in AD.

### TREM2 genotype dependence of differential gene expression in excitatory neurons with increasing β-amyloid

*TREM2var* increase risks of cognitive impairment [25]. We previously provided evidence for greater loss of vulnerable neurons with the *R47H TREM2var* [8]. Here we hypothesised that, while mediated through functional differences in microglia, risk variants would be associated with more deleterious neuronal transcriptional responses to increasing tissue β-amyloid than is found with *CV*. To explore this hypothesis, we comprehensively assessed differential gene expression with increasing β-amyloid in both excitatory and inhibitory neuronal subtypes (**Supp. Fig. 3a, d, e**). As expected, given the lower number of samples, the proportions of differentially expressed genes relative to the total expressed genes generally were lower for *TREM2var* (**Supp. Fig. 3d, e**). We began by exploring differentially expressed pathways in L2-3 and L4-5 excitatory neurons where differential gene expression was the highest across genotypes.

L2-3-*CUX2* neurons from all genotypes showed similar ratios of genes up- and down-regulated (*CV*, R_g_= 1.1 (598 DEG); *R62H*, R_g_ 1.0 (316 DEG); *R47H,* R_g_= 0.5 (254 DEG)) with increasing β-amyloid however reduced DGE in *TREM2var* (**Supp. Fig. 3d; Supp. Table 7**). We found greater numbers of differentially expressed gene pathways enriched in the *TREM2var* than in the *CV* samples (**Supp. Table 23**). There was an upregulation of transcripts encoding for ribosome subunits and involved in pathways such as ‘cytoplasmic translation’ and ‘cytosolic large ribosomal subunit’ in *CV* and *R62H*. These neurons also were enriched for mitochondrial genes (*NDUFA4, COX5B*) and pathways (‘Mitochondrial electron transport chain, cytochrome C to oxygen’ and ‘oxidative phosphorylation’). Several plasticity pathways were upregulated such as those for ‘axon guidance’ (*SEMA3C*) and ‘post-synaptic density’ (*GRIA1*, *GRIN1*) (**Fig. 5**). *R62H* L2-3-*CUX2* neurons also upregulated translation, axonogenesis (*SEMA3D*) and post-synaptic density (*SYT11*) pathways. By contrast, *R47H* neurons did not upregulate translation, mitochondrial or plasticity pathways. Interestingly, both *TREM2var* samples upregulated expression of *RTN4*, in the context of ‘negative regulation of β-amyloid formation’ pathways. Upregulated *RTN4* expression may enhance disease progression [32]. *R47H* also had an upregulation of *ATP1A3* involved in ‘β-amyloid binding’ and leads to amyloid-induced neuronal degeneration [44]. Amongst the small numbers of pathways downregulated were ‘exocytic vesicle membrane’ in *CV*, ‘potassium channel activity’ in *R62H* and a number of synaptic activity-related pathways in *R47H* samples (**Fig. 5**).

**Fig. 5.**
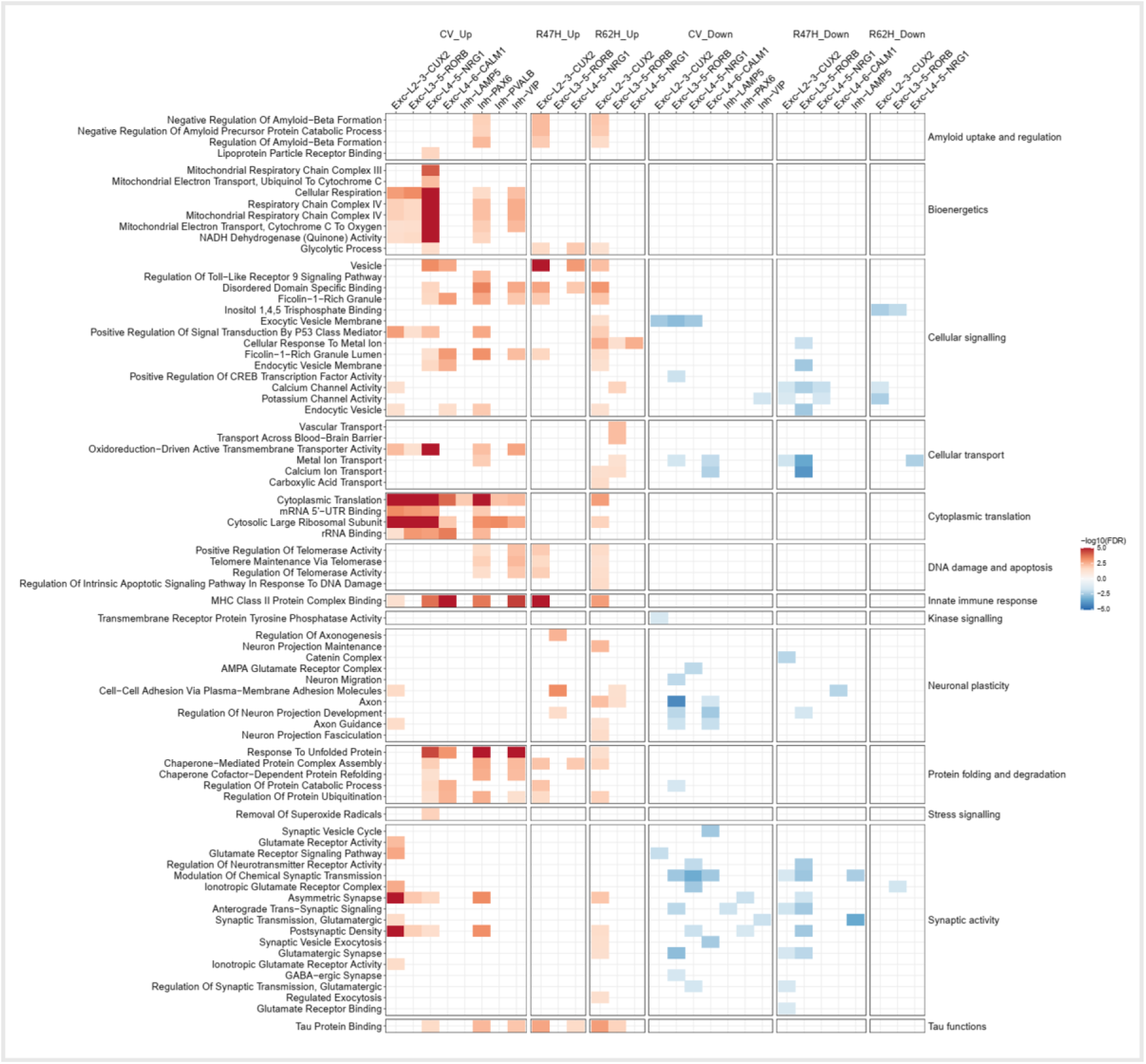
Differential gene expression of excitatory and inhibitory neuronal subtypes with different TREM2 variants. CV neurons had a range of upregulated pathways including those associated with ‘cytoplasmic translation’ and ‘response to unfolded protein’ across excitatory and inhibitory subtypes. TREM2var cases had fewer upregulated pathways. All genotypes had fewer down-regulated pathways with synaptic activity pathways being the most affected in CV and R47H cases.

In contrast to the L2-3 neurons, *CV* and *R62H* L4-5-*NRG1* neurons showed similar numbers of genes up- and down-regulated (*CV,* R_g_= 1.3 (913 DEG); *R62H*, R_g_= 0.8 (24 DEG)) while neurons in *R47H* tissue showed more down-regulated genes (R_g_= 0.2 (227 DEG)) with increasing β-amyloid (**Supp. Fig. 3d; Supp. Table 9**). *CV* L4-5-*NRG1* neurons showed the greatest differential expression of genes with increasing β-amyloid. *CV* L4-5-*NRG1* neurons upregulated genes in pathways with potentially neuroprotective functions, such as enhancing protein degradation via ‘response to unfolded protein’ (*DNAJA1, DNAJB1*) and ‘regulation of protein ubiquitination’ (*UBB*) (**Supp. Table 25**). Similar to L2-3-*CUX2* neurons, translation, mitochondrial and synaptic activity pathways were also up-regulated in the *CV* group but not in *TREM2var* (**Fig. 5**). *CV* and *R47H* L4-5-*NRG1* neurons upregulated transcripts (*ATP1A3, CLU*) associated with ‘β-amyloid binding’ but few pathways were upregulated in *R62H. CV* L4-5-*NRG1* neurons mainly down-regulated pathways associated with synaptic transmission such as those for the ‘AMPA glutamate receptor complex’ *(GRIA3, CACN8)* and ‘modulation of chemical synaptic transmission (*DLGAP4, SYN3, OPHN1*). *TREM2var* had few downregulated pathways including downregulated ‘calcium’ and ‘potassium channel activity’ in *R47H* and ‘metal ion transport’ (**Fig. 5**).

These results suggest that with increasing β-amyloid in AD there is greater upregulation of genes involved in potentially neuroprotective pathways in excitatory neurons in*TREM2 CV* samples relative to those from risk variant carriers. *TREM2var* cases were distinguished by a potentially maladaptive upregulation of pathways involved in β-amyloid formation as a consequence of pathologically the increased activity or stress associated with *TREM2var* heterozygosity.

### TREM2 genotype dependence of differential gene expression of inhibitory neurons with increasing β-amyloid

We found greater differential gene expression in interneurons in the CV group than in the *TREM2var* cases (**Supp. Tables 13-17**). *PAX6* inhibitory neurons had proportionally more genes upregulated than other inhibitory neurons in *CV* (*PAX6*, R_g_=0.6 (142 DEG); *VIP*, R_g_= 0.8 (80 DEG); *LAMP5*, R_g_= 0.6 (31 DEG); *PVALB*, R_g_= 0.6 (22 DEG); *SST*, R_g_= 0.3 (4 DEG)). Fewer differentially expressed genes were found in *TREM2var* genotypes: <7 DEGs in *PVALB*, *SST, PAX6* and *VIP* neurons with both *TREM2var*. *LAMP5 R47H* inhibitory neurons had the most differential expression (R_g_= 0.2 (14 DEG)) of the *TREM2var* with none in *R62H LAMP5* neurons.

All *CV* inhibitory subtypes were enriched in transcripts associated with ‘cytoplasmic translation’, similar to the excitatory neurons (**Supp. Table 27-30**). *CV VIP* and *PAX6* neurons were enriched for many of the same pathways (e.g., ‘cellular respiration’, ‘response to unfolded protein’) (**Fig. 5**). *PAX6* interneurons upregulated transcripts associated with ‘regulation of β-amyloid formation’ (*RTN1, ROCK2, RTN4*) and the ‘post-synaptic density’ (*DISC1, MAGI2*). However, *PAX6* neurons also downregulated a number of transcripts associated with ‘asymmetric synapse’ (*SPOCK1, ADGRA1)*.

We found few significantly up or downregulated pathways in *TREM2var* inhibitory neurons. *LAMP5* interneurons had a downregulation of transcripts (*UNC13A, GRID1*) involved in the ‘synaptic transmission, glutamatergic’ pathway in the *R47H* carriers (**Fig. 5**).

Together, these results describe upregulation of several potentially adaptive pathways in inhibitory neuronal sub-types related to processing of abnormal proteins, bioenergetics, and synaptic activity in *CV* relative to *TREM2var* tissues (**Fig. 5**). *R47H* inhibitory *LAMP5* neurons down regulated genes for synaptic transmission, potentially permissive for greater excitatory neuronal disinhibition. *R62H* interneurons were associated with few significant DEG or pathways.

### Bulk cortical proteomics extends transcriptomics and suggests genotype-dependent differences in neuronal stress and death pathway activation

Proteomics provides a complementary approach to characterisation of bulk cortical tissue responses to increasing β-amyloid. Numbers of significantly differentially expressed proteins with greater β-amyloid load differed by genotype: *R47H* cases had 28.3% (908) of total detected proteins (p<0.05, unadjusted), *R62H* cases had 4.7% (150) and *CV* cases had 5.9% (189) differentially expressed proteins (DEPs) (**Supp. Table 35**).

Interpretation of the relative numbers of DEPs is confounded by the influence of differences in sample numbers as well as the effects of differences in genotype (*CV>R62H>R47H*). However, differences in the nature of DEPs may provide pathological insights. After adjustment for multiple comparisons, we found evidence of few impacted pathways in *CV* cases. *R62H* and *R47H* tissue responses were different apart from the shared upregulation of ‘glutathione transferase activity’, a stress response [1]. *R47H* cases had an upregulation of proteins (FKBP1A, GSN) involved in ‘amyloid fibril formation’. Downregulated proteins in *R47H* included those involved in synaptic transmission pathways such as ‘syntaxin binding’ (SNAP25, VAMP1/2) and ‘synaptic vesicle budding’ (PICALM, SNAP91). *R62H* cases, in which neurons as previously described upregulated the expression of proteins associated with the regulation of protein ubiquitination, had an upregulation of proteins associated with ‘ubiquitin conjugating enzyme activity’ (UBE2D2, UBE2Z) and ‘deubiquitinase activity’ (CYLD, USP47, USP31). *R62H* cases had a downregulation of proteins associated with ‘β-amyloid binding’ (APOA1, CST3), functional effects of which may exacerbate β-amyloid accumulation [10, 12].

We next performed a direct, exploratory comparison of differently expressed transcriptional and proteomic pathways using a more permissive threshold based on nominal rather than adjusted *p* values (**Supp. Fig. 4, Supp. Tables 36-38**). Most strikingly, pathways upregulated at the transcript level in CV astrocytes and neurons were broadly downregulated at the protein level in *TREM2var* including astrocytic and neuronal mitochondrial pathways and ‘cellular translation’ pathways in neurons. Upregulated amyloid-related transcriptomics pathways in *TREM2var* astrocytes such as ‘response to β-amyloid’ and ‘regulation of β-amyloid formation’ showed consistent downregulation at the protein level in *CV* and *R47H* cases, although they showed consistent upregulation in the *R62H* cases.

‘Response to unfolded’ protein was upregulated at the transcript and protein levels in astrocytes and neurons and ‘regulation of amyloid fibril formation’ was upregulated at both levels in neurons in *CV.* Proteins involved in the ‘synaptic transmission, glutamatergic’ pathway, whilst transcriptionally upregulated, were downregulated in *CV* neurons. Most neuroplasticity pathways transcriptomically downregulated (e.g., ‘axonogenesis’ ‘glutamate receptor signalling pathway’, ‘modulation of chemical synaptic transmission’) were also downregulated at the protein level in *CV* neurons.

In *R47H* astrocytes, whilst β-amyloid response pathways (’regulation of β-amyloid formation’) were upregulated transcriptomically, associated proteins were downregulated. The ‘response to β-amyloid’ was down regulated at both RNA and protein level in *R47H* neurons as were neuroplasticity pathways (‘modulation of chemical synaptic transmission’, ‘regulation of synaptic transmission, glutamatergic’).

*R62H* astrocytes upregulated expression of the ‘regulation of β-amyloid formation’ pathway at both RNA and protein levels. However, they had discordant changes in neuroplasticity support pathways such as ‘anterograde trans-synaptic signalling’, ‘glutamate receptor signalling’ pathways which, although downregulated transcriptomically, were upregulated at the protein level. Conversely, plasticity pathways associated with synaptic vesicle function (‘synaptic vesicle endocytosis’, ‘synaptic vesicle exocytosis’) were upregulated at the RNA level although associated proteins were downregulated. *R62H* cases upregulated ‘calcium ion transport’ and related pathways at both the astrocyte and neuronal transcriptomic and protein level, potentially consistent with neuronal hyperactivity and its regulation [49], which were mostly downregulated in *R47H* and unaffected in *CV*.

Together, these proteomic data extend transcriptomic observations and highlight genotype-specific evidence for adaptive glial-neuronal adaptive plasticity particularly in *CV* tissues and increased neuronal stress responses and β-amyloid production with *TREM2var* with increasing β-amyloid.

## Discussion

By contrasting the cellular pathology associated with heterozygosity for the *R47H* and *R62H* risk variants with that of the common allele, our study has provided new insights into the role of *TREM2* in AD. We found that both risk variants were associated with higher β-amyloid load and, for the first time in humans, showed that this was dependent on the *CD33* genotype. This was reflected as an increased density of compact rather than diffuse plaques, suggesting that microglial plaque compaction functions are preserved, in contrast to earlier observations with microglia that are *TREM2 R47H* homozygous [3].

The rs3865444 *CD33* polymorphism is associated with reduced expression of the receptor. *CD33* knockout in the 5XFAD model showed TREM2-dependent attenuation of β-amyloid pathology deposition, providing evidence for epistasis [17]. We found evidence for a similar *CD33-TREM2* epistasis in human AD with relative normalisation of differences in β-amyloid load particularly in *R47H* cases mainly driven by a reduction in plaque size in those carrying the protective *CD33var*. Microglial transcriptomic responses to β-amyloid were found in *R62H* cases in the context of the protective *CD33* genotype. This is consistent with the short CD33 isoform being a gain-of-function variant [5] which can compensate for *TREM2var*-induced reduction in glial activation by rebalancing immunoreceptor tyrosine-based activation (ITAM) and inhibition (ITIM) in the microglia [47]. Functional effects were reflected across the broad range of microglial gene expression pathways regulated by *TREM2* signalling, including phagocytosis, inflammatory response and lipid metabolism [60]. Differences in pathways significantly modulated also highlighted differences in signalling pathologies across these pathways for the risk genotypes. Consistent with Griciuc et al. [17], we found that the rescue of microglial transcriptomic changes in the *TREM2var* with the strongest effect on microglial gene expression (*R47H*; **Supplementary Figure 3**), was not present in the presence of *CD33var*. However, we employed a conservative analysis approach, such as using a mixed model with linear regression to avoid false positives and a correction for pathology load, to more accurately inform on the effects of effects of genotype at the level of microglial function. This approach, together with the low sample number in the *R47H-CD33 CV* group, may explain the lack of significant effect.

Functional changes with *TREM2 KO* or the *R47H TREM2* allelic variant have been characterised in microglia *in vitro* and in animal models [14, 43, 48, 61]. β-amyloid clearance is reduced in *TREM2 KO* mice [14] with increased expression of stress-associated autophagocytic vesicles, impaired microglial lipid metabolism and decreased survival [43, 48, 61]. Controlling for *CD33* genotype, we found that microglial transcriptional responses to increasing β-amyloid were lower for *TREM2var*, particularly for the *R47H* variant. Whilst there were fewer *R47H* samples, a statistical power calculation suggested that, at our actual R47H sample size, 34.8% of *CV*-associated genes would be detectable at 0.8 power if they had equivalent logFC. The attenuated response in *R47H* therefore reflects a genuine biological dampening of the amyloid-beta associated microglial transcriptional response, consistent with the known role of *TREM2 R47H* in impairing microglial response to amyloid. *R62H* microglial signatures were distinguished from those of *R47H* by upregulation of genes associated with phagocytosis and β-amyloid binding. This illustrates that differences in TREM2 pathology are associated with gain, as well as loss of function.

Fewer studies have described the secondary consequences of *TREM2 KO* or *TREM2var* expression on astrocytes and neurons. Initial studies of astrocytes in mouse *TREM2 KO* models suggested few effects on their transcriptomic signatures [33], although reactive astrocytes in the *TREM2 KO* [56] may engulf synapses [22]. We found substantial *TREM2* genotype associated differences in astrocytes with AD. *R62H* astrocytes differentially expressed greater numbers of genes than for the common allele, including those involved in NF-kappaB signalling. This analysis also suggested differences between astrocyte responses associated with the three *TREM2* genotypes. For example, we found evidence for significant up regulation of the gene expression pathways related to protein clearance (e.g., ubiquitination pathways) and neuronal plasticity (e.g., neuronal growth pathways) with the CV genotype. consistent with work describing a role for astrocytic *TREM2* in neuro-repair in a stroke model [64]. By contrast, *TREM2var* astrocytes downregulated the neuronal plasticity (*R47H*) and glutamatergic synapse pathways (*R62H*). The greater neurodegeneration that we have reported with *TREM2var* pathology [8] thus is a consequence of reduced neurotrophic support as well as increases in locally neurotoxic factors (e.g., β-amyloid). Our work suggests that the “loss of function” mechanistic hypothesis for TREM2-related pathology, which is supported by observations such as the association of *TREM2* with synaptic “eat me” signals, needs to be extended to include secondary effects of astrocytes [54]. These observations highlight that, while *TREM2* pathology is mediated through microglia primarily expressing the variant gene amongst brain cells, expression of the pathology is determined by multi-cellular interactions.

Evidence for neuronal contributions to the pathology also was found. Differential gene expression with increasing β-amyloid suggested upregulation of β-amyloid processing pathways in *TREM2var* excitatory neurons and lower enrichment for adaptive pathways expressed in *CV* inhibitory neurons for both *TREM2var.* Exploratory bulk tissue proteomics supported these observations with evidence for adaptive plasticity and protein degradation in response to β-amyloid pathology in *CV* tissue. This was not found for the *TREM2var* although they showed evidence for altered β-amyloid handling and synaptic plasticity. These results thus suggest that the increased β-amyloid load with *TREM2var* is a consequence of reduced processing of β-amyloid with neuronal stress as well as altered glial processing of amyloid. The reduction or absence of appropriate β-amyloid clearance responses in *TREM2var* microglia along with the potential reduction of astroglial neuronal support and direct toxic effects associated with relatively increased β-amyloid together all likely contribute to the increased neuronal vulnerability with *TREM2var*.

Previous reports that the *R47H* mutation reduces *TREM2* affinity for β-amyloid and lipid ligands and reduces clearance *in vitro* have suggested that *TREM2var* exert effects through lack of amyloid clearance by microglia with AD [65, 67]. Our results suggest the pathogenesis is more complex. While we showed that *R47H* microglia had a reduced response to β-amyloid as reported by others, we also demonstrated that *R62H* microglia have some increased responses to β-amyloid including an increased activation morphology and an upregulation of phagocytosis pathway genes which may contribute to the increased plaque compaction. An earlier report suggested increased plaque seeding with *TREM2var* or *KO* [14, 45]. Our work suggests that an alternative explanation for increased amyloid accumulation is downregulation of degradation mechanisms in the *TREM2var*. Moreover, we have defined additional potential contributions to the accelerated progression of pathology from astrocytes and neurons Therefore, neuronal responses should also be expected (and explored) with therapeutic *TREM2*-activator strategies [34].

A major strength of our report is the joint characterisation of glial and neuronal transcriptional pathology in healthy and diseased human cortical tissue across *TREM2* genotypes. However, we recognise that our work also has several limitations. Availability of tissue from the rare *TREM2var* donors limited the numbers of brains and regions available. Tissue with earlier pathology (Braak stages III-IV) and NDC (Braak 0-II) [6] were especially limited. We sought to mitigate bias towards late-stage AD by sampling regions with lower (SOM) and moderate (MTG) pathology and through regression against β-amyloid in the same region (contralateral hemisphere) as used for transcriptomics, allowing the DGE to be in the context of local pathology. The scarcity of *TREM2var* tissue meant that there was some imbalance between the groups in terms of the proportion of *CD33var* and *APOE4* carriers, *post-mortem* interval and RNA integrity. We sought to mitigate this by including *APOE4* as a covariate in the transcriptomic analysis and by using a joint linear regression model however this may explain the lack of *CD33*-dependent effects on amyloid pathology in *R62H* and differential gene expression in *R47H*. However, statistical power calculations suggested that our data was biologically meaningful. The joint model, incorporating all data into a single design matrix, mitigates biases arising from unbalanced AD/control sample sizes across subgroups—as would confound genetically stratified analyses. As we were limited to relatively small amounts of frozen tissue, we employed snRNAseq which is sparse and biased compared to whole cell transcriptomics and with a lower representation of glial inflammatory responses [2, 57], which may limit overlap with proteins. Contralateral homologous fixed sections were used for histopathology, rather than adjacent sections from the tissue sections used for transcriptomics. However, while this may reduce the analytical power by increasing variance, we do not believe it introduces major bias because of the approximately bilateral symmetry of AD pathology [41]. snRNAseq and *post-mortem* tissue artifacts may limit accurate characterisation particularly of cellular inflammatory responses and depend on the tissue storage conditions [57, 68]. Whilst we characterised the global β-amyloid and quantified the subtypes of β-amyloid plaques, we did not characterise any potential differences in β-amyloid molecular aggregates arising from differences in microglial uptake and degradation [55]. Furthermore, spatially-dependent differences in glial responses and microglial-astrocytic interactions to β-amyloid (e.g., periplaque vs distant from plaques) were not characterised. Understanding molecular β-amyloid species and the local impact of plaques would allow for more targeted thereapeutic approaches.

Together, these results highlight differences in molecular pathology between the *TREM2var* risk variants. Although both variants increase AD risk, *R47H* confers a substantially greater risk than *R62H*. We therefore investigated whether the two variants have distinct biological consequences. While both variants are associated with altered microglial function, their downstream cellular responses differ, and we propose that these differences underlie the divergent levels of risk. Our results demonstrate how their impact on AD risk may be mediated by secondary effects on astroglial and neuronal function, as well as the primary microglial functional pathology. The evidence for contributions to β-amyloid clearance by astrocytes and genotype-associated differences in microglial activation that we have described may alter soluble amyloid oligomer and fibril levels. Quantitative characterisations of these across *TREM2* genotypes should be a priority for understanding differences in AD risk conferred [7]. While *TREM2* activators are promising to explore, demonstration of strong epistasis between *TREM2* and *CD33* with AD should be considered in *TREM2* agonist trials and may also support the therapeutic potential of modulators of CD33 expression. The latter could offer some advantages given the accumulated clinical experience with some agents already [63].

## Author contributions

NF led the snRNAseq and proteomics analysis. NW led the IHC and snRNAseq data generation. VC conducted IMC analysis and IHC microglia analysis. SB conducted IHC plaque analysis. HW, BC and WW conducted the LCMS. ST contributed to pathway analysis. BA, MT and JTM processed raw RNAseq data. CK established snRNAseq pipeline. AMG, KD and AS conducted snRNAseq. ES conducted genotyping analysis. RY conducted protein extractions. DC, EA and MP conducted histological staining. WS and JH conducted genotyping. JSJ and PMM conducted study design, staff supervision, analysis and interpretation of data and funding. JSJ wrote the first draft of the manuscript. PMM joined in drafting and editing the manuscript and arranging funding for the work. All authors contributed, reviewed and approved the manuscript.

## Acknowledgements

We thank the donors and their families for the use of human brain tissue in this study and the UK brain bank staff for making it available. Tissue samples were provided by the London Neurodegenerative Diseases Brain Bank at King’s College London. The brain bank receives funding from the UK Medical Research Council and as part of the Brains for Dementia Research programme, jointly funded by Alzheimer’s Research UK and the Alzheimer’s Society. Tissue for this study was provided by the Newcastle Brain Tissue Resource which is founded in party by a grant from the UK Medical Research Council (G0400074), by NIHR Newcastle Biomedical Research Centre and Unit awarded to the Newcastle upon Tyne NHS Foundation Trust and Newcastle University, and as part of the Brains for Dementia Research Programme jointly funded by Alzheimer’s Research UK and Alzheimer’s Society. In addition, tissue was provided by the Queen’s Square Brain Bank, UCL. Tissue samples were supplied by The Manchester Brain Bank, which is part of the Brains for Dementia Research programme, jointly funded by Alzheimer’s Research UK and Alzheimer’s Society. We acknowledge the Oxford Brain Bank, supported by the Medical Research Council (MRC), the NIHR Oxford Biomedical Research Centre and the Brains for Dementia Research programme, jointly funded by Alzheimer’s Research UK and Alzheimer’s Society. Tissue samples and associated clinical and neuropathological data were supplied by Parkinson’s UK Brain Bank at Imperial, funded by Parkinson’s UK, a charity registered in England and Wales (258197) and in Scotland (SC037554). We are grateful to Diana P. Benitez for her support in the human tissue management. Figure 1 was created with BioRender.com.

Infrastructure, including particularly the LMS/NIHR Imperial Biomedical Research Centre Flow Cytometry Facility and the Imperial BRC Genomics Facility, was supported by the National Institute for Health Research (NIHR) Biomedical Research Centre (BRC). ST was supported by an “Ambizione” career development grant (223414) and an “Early Postdoc Mobility” scholarship (191446) from the Swiss National Science Foundation and a “Clinical Medicine Plus” scholarship from the Prof Dr Max Cloëtta Foundation (Zurich, Switzerland). JH acknowledges support from the Dolby Foundation and the UK DRI. PMM acknowledges generous personal support from the Edmond J Safra Foundation and Lily Safra and an NIHR Senior Investigator Award. This work was supported by a grant to PMM from the UK Dementia Research Institute, which receives its funding from UK DRI Ltd., funded by the UK Medical Research Council, Alzheimer’s Society, and Alzheimer’s Research UK. JSJ was supported by funding from Alzheimer’s Society (grant number 599 (AS-DRL-22-008)). The study is an output from the UK Dementia Institute Multi-omics Atlas Project for Alzheimer’s Disease (MAP-AD; map-ad.org).

